# Distinct Diurnal and Day of Week Online Search Patterns Related to Common Eye Conditions

**DOI:** 10.1101/2020.07.04.20146449

**Authors:** Michael S. Deiner, Stephen D. McLeod, Julie M. Schallhorn, James Chodosh, Daniel H. Hwang, Thomas M. Lietman, Travis C. Porco

**Affiliations:** F. I. Proctor Foundation, University of California, San Francisco, San Francisco, California; Department of Ophthalmology, University of California, San Francisco, San Francisco, California; Department of Ophthalmology, Massachusetts Eye and Ear, Harvard Medical School, Boston, Massachusetts; The Nueva School, San Mateo, California; F. I. Proctor Foundation, University of California, San Francisco, San Francisco, California; Department of Ophthalmology, University of California, San Francisco, San Francisco, California; Department of Epidemiology and Biostatistics, Global Health Sciences, University of California, San Francisco, San Francisco, California

## Abstract

**Importance:** Studies suggest diurnal patterns of some eye conditions. Leveraging new information sources such as online search data to learn more about such patterns could improve understanding of patient eye-related conditions and well-being and improve timing of clinical and remote eye care.

**Objective:** To investigate our hypothesis that the public is likely to consistently search about different eye conditions at different hours of the day or days of week, we conducted an observational study using search data for terms related to eye conditions such as conjunctivitis. We asked if search volumes reflected diurnal or day-of-week patterns and if those patterns were distinct from each other.

**Design:** Hourly search data for eye-related and control search terms for 2018 were analyzed and compared.

**Setting:** Data from 10 USA states.

**Exposure:** Internet search.

**Participants:** Populations that searched Google’s search engine using our chosen study terms.

**Main Outcome Measures:** Cyclical hourly and weekly online search patterns.

**Results:** Distinct diurnal (p<0.001 for all search terms) and day-of-week search patterns for eye-related terms were observed but with differing peak time periods and cyclic strengths. Some diurnal patterns represented reported clinical patterns. Of the eye related terms, “conjunctivitis” and “pink eye” had the strongest diurnal cyclic patterns based on peak-to-trough ratios. Stronger signal was restricted to and peaked in mornings, and amplitude was higher on weekdays. In contrast, “dry eyes” had a higher amplitude diurnal pattern on weekends, with stronger signal occurring over a broader evening to morning period and peaking in early morning.

**Conclusions and Relevance:** The frequency of online searches for various eye conditions can show cyclic patterns according to time of day or week. Further studies to understand the reasons for these variations may help supplement current clinical understanding of eye symptom presentation and improve the timeliness of patient messaging and care interventions.

**Key Points:** *Question:* Do online public search engine queries for different eye-health terms follow hourly or daily patterns and do the patterns differ from each other or reflect what is known clinically?

*Findings:* Unique hourly and day of week eye health related search patterns appear diurnal and can reflect what has been observed clinically.

*Meaning:* Online search data may reflect timing of eye conditions and could improve clinical understanding of eye-related symptom occurrence, including outside of clinics. Knowing precisely when patient’s eye condition interests increase holds promise -for example to optimize timing and availability of local or remote eye care resources.

## Introduction

Clinical study has identified cyclic occurrence of health conditions in humans, including eye-related conditions, and may facilitate chronopreventive and chronotherapeutic care^1–8^. Online search behavior regarding disease symptoms has been shown to reflect seasonal and diurnal clinical patterns as well as aspects of disease not typically observed in clinics at all (for example, coronary heart disease and depression)^9,10^. Online search or social media data can reflect seasonal clinical eye disease patterns and conjunctivitis epidemics^11–15^. Here we tested the hypothesis that the public is likely to search about different aspects of eye-health at different hours of the day or days of week. Specifically, we conducted an observational study investigating if US hourly online search data for terms related to conjunctivitis or other common eye conditions and treatments reflected diurnal or day-of-week patterns and if those patterns were distinct from each other.

## Methods

### Google search data

For conjunctivitis and for comparison for other common eye conditions and treatments, we selected the search terms “conjunctivitis”, “blurry eyes”, “cataracts”, “pink eye”, “dry eyes”, “watery eyes”, “glaucoma”, “contact lenses”, “visine”, “lasik”. We also included a positive control term that would likely exhibit hourly and day of week variation (“drunk”). Relative hourly search frequency volume (RSV) data for these terms for the year 2018 from the 10 most-populous US states (CA, FL, GA, IL, MI, NC, NY, OH, PA, TX) were downloaded in April 2019 from Google Trends^9,10^. Universal Coordinated Times were adjusted to the predominant time zone for each state (only FL, MI and TX include multiple time zones). The resulting time series represented RSV for a given time period and term. Data for all states was combined and mean hourly results for each term were then normalized for visual comparison in polar plots (R package ‘ggplot’).

### Diurnal and day of week analysis and comparison of cyclic strength and peak times

Using the RSV for each search term as an outcome variable, we conducted trigonometric regression adjusting for trend, as follows. We adjusted for overall trend using sixth-order orthogonal polynomials in the number of days since 1 Jan. Diurnal effects were modeled by terms of the form sin *nωt* and cos *nωt*, where *ω=2π/24, t* is the time measured on a 24 hour clock, and *n*=1,…,4. We estimated separate diurnal effects for weekend days and for weekdays. Outliers and localized (nonperiodic) departures were modeled by terms of the form 1_*x*∈*[16m,16(m+1)-1)*_, where *m*=0,1,… and *x* is the number of hours elapsed since midnight, 1 Jan. Additional terms of similar form were added to represent weeks. Inclusion of these terms was determined by a cross-validated LASSO procedure. Following model selection, ordinary least squares estimation was used to estimate the trend, outlier, and trigonometric coefficients. From the trigonometric coefficients and intercept, we estimated the circular median occurrence time and the peak-to-trough ratio (in a similar fashion as our previous analyses and using the R Package ‘circular’)^12–14^. Because diurnal and day of week occurrence data are angular data, we used the circular median time to summarize the central tendency; the circular median reflects the peak occurrence (when the data are approximately unimodal). The peak-to-trough ratio represents cyclic strength, or amplitude. Standard errors and *P*-values were determined using time series bootstrap. *P*-values less than 0.001 were considered significant. UCSF IRB approval #14-14743 was obtained for this study.

## Results

Overall, we found each search term exhibited diurnal patterns of search interest (p < 0.001 for all terms). However, cyclic strength and central tendency differed between search terms, as described below.

### Hourly and weekly patterns

To visualize cyclic diurnal patterns for each term, mean RSV at each time of day is represented in polar 24-hour plots in Figure 1. To allow visual comparison between terms, results for each term are normalized so that the hours with the least RSV interest are closest to center, while hours of higher RSV interest are further from center. Note that despite most terms exhibiting diurnal patterns, scale bars in Figure 1 indicate not all terms exhibited similar diurnal strength. In Figure 2 cyclic diurnal patterns for each term on each day of the week were plotted. These plots suggested some terms had diurnal cyclic features that varied between weekdays and weekends.

**Figure 1.**
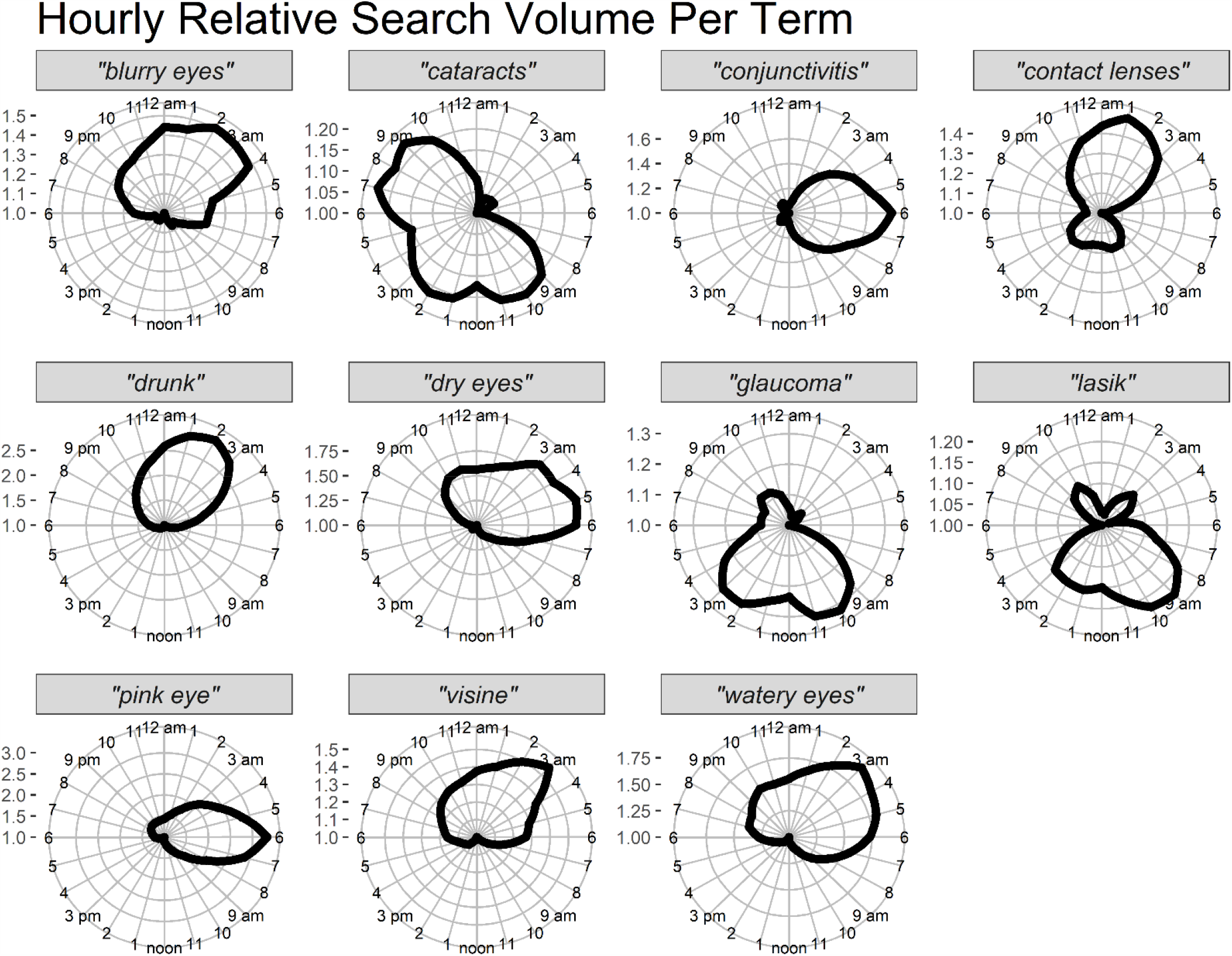
Average hourly pattern from 2018 for 10 USA States combined. For each term the RSV is shown, with lowest relative hourly interest plotted near center and higher value hours plotted further from center. In order to optimally demonstrate cyclic patterns per terms, RSVs for each term were normalized by dividing the mean per each hour per term by the value of the hour having the smallest mean value. The hours with the least RSV are closest to center with value of 1.0, while hours of higher relative search interest are further from center. All terms were shown to have statistically significant diurnal patterns (for all terms shown, please see Table 1 for P values along with hourly mean peak values and bootstrapped CI and statistical significance). Please note however, that the scale bars for each term in Figure 1 indicate not all terms exhibited similar diurnal strength and that the plots do not represent total search interest for one term vs. another.

**Figure 2.**
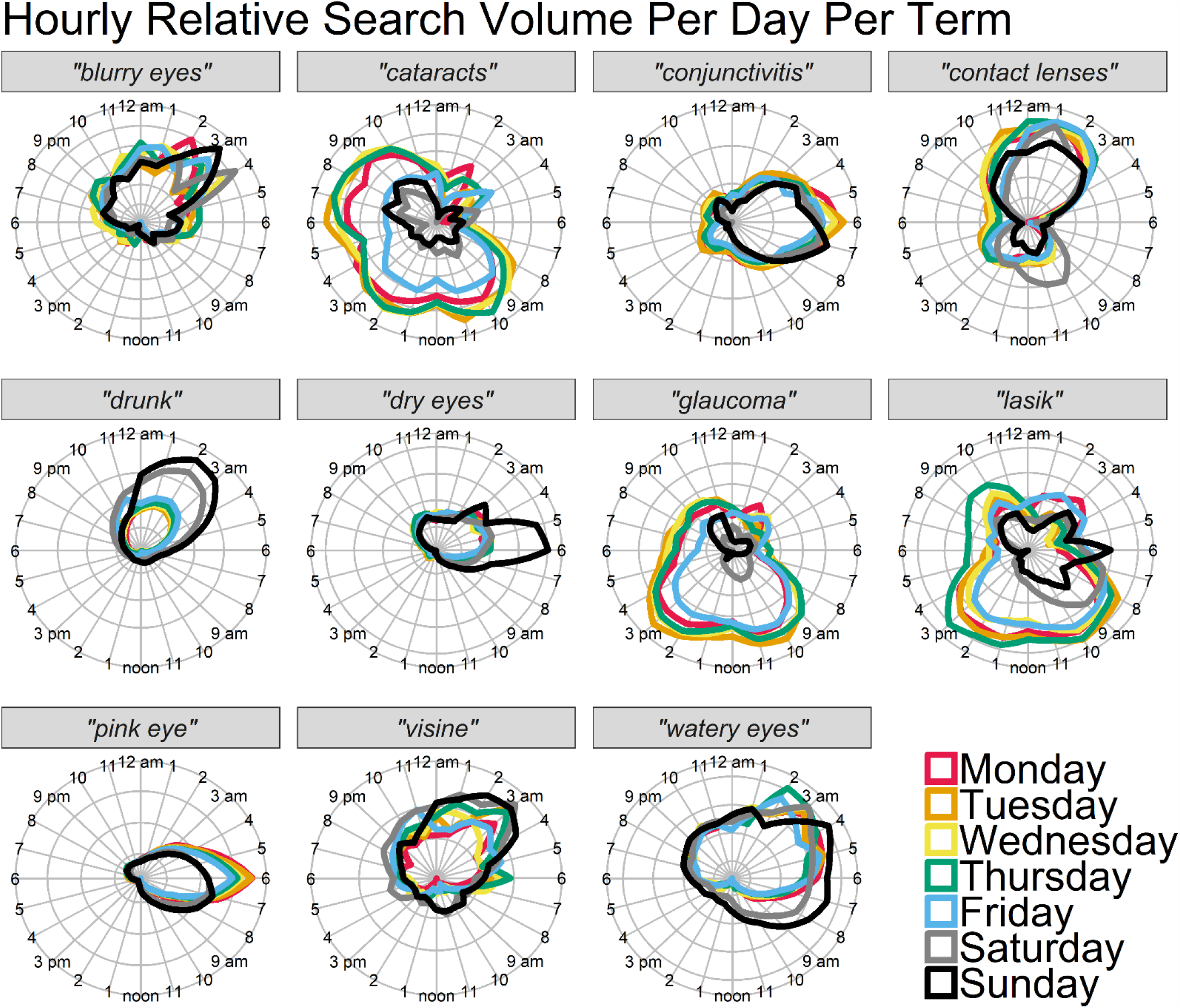
Average hourly pattern per weekday from 2018 for 10 USA States combined. For each term the RSV is shown per day of week, with lowest relative hourly interest plotted near center and higher value hours plotted further from center. Values have been normalized as in Figure 1 and do not represent total search interest for one term vs. another - but relative amount of search interest between days for an individual term is visually represented. Terms shown to have statistically significant day of week patterns, mean peak day values and other day-of-week characteristics for all terms are shown in Table 1.

### Statistical analysis of patterns

Following smoothing and detrending, the resulting dataset was used for all subsequent statistical analyses. Results for all terms are presented in Table 1. Column 2 provides peak-to-trough ratios. Columns 3-6 provide weekday and weekend circular median times as well as differences between weekday and weekend for the peak times and for amplitude. For all terms, diurnal patterns were significant on weekdays and on weekends (p <0.001 for all terms). Of the eye-related terms, “conjunctivitis” and “pink eye” had the strongest diurnal cyclic patterns based on peak-to-trough ratios, with stronger signal restricted to a narrow time window and peaking all mornings within the same 1 hour period. These showed a higher amplitude on weekdays (see Table 1, columns 2, 5, 6; Figure 2). In contrast, “dry eyes” exhibited a stronger diurnal pattern on weekends, with stronger signal occurring over a broader evening to morning time window, peaking in early morning (see Table 1, column 5, Figure 2). For other terms, larger differences between weekday and weekend were seen. For example, “glaucoma” peak times were over 11 hours apart, exhibiting much stronger amplitude on weekdays and occurring over a very broad period. Similar results occurred for “cataracts” and “lasik”. The control term “drunk” exhibited the highest peak-to-trough ratio with a much stronger amplitude on weekends.

**Table 1.** Cyclical diurnal or day of week characteristics of relative search values. Column 1: Search terms; Column 2: “Peak to Trough Ratio” for all days combined per term; a larger average daily peak-to-trough value indicates a more pronounced diurnal pattern. For the remaining columns, weekdays were assessed separately from weekend day and compared. The average filtered and detrended circular median time (and 95% CI) are shown for each term for weekdays (Column 3: “Weekday Circular Median Time”) and for weekend days (Column 4: “Weekend Circular Median Time”). The asterisks indicate evidence that the diurnality seen is unlikely to result from chance alone (p < 0.001); Column 5: “Circular Median Difference” categorizes the time differences between the weekday and weekend circular median times, for each term; Column 6: “Weekend Amplitude Difference” allows a comparison of weekday to weekend diurnal cycle amplitudes (i.e. a comparison of cyclic strength), providing the average difference (and 95% CI) between weekday vs. weekend amplitudes. Negative values indicate stronger weekday cyclic strength and positive values indicate stronger weekend cyclic strength. Values further from 0 indicate a larger difference between weekdays and weekend days.

| Search Term | Peak to Trough Ratio | Weekday Circular Median Time | Weekend Circular Median Time | Circular Median Difference | Weekend Amplitude Difference |
| --- | --- | --- | --- | --- | --- |
| *“blurry eyes”* | 1.37<br>(1.29, 1.45) | 01:04*<br>(00:46, 01:20) | 01:27*<br>(00:58, 01:53) | <1 hour | -0.11<br>(-0.19, -0.04) |
| *“cataracts”* | 1.27<br>(1.23, 1.31) | 15:18*<br>(15:03, 15:32) | 18:51*<br>(17:33, 20:26) | >3 hours | -2.1<br>(-2.3, -1.9) |
| *“conjunctivitis”* | 1.60<br>(1.56, 1.65) | 06:13*<br>(06:05, 06:20) | 05:49*<br>(05:41, 05:58) | <1 hour | -1.9<br>(-2.2, -1.7) |
| *“contact lenses”* | 1.34<br>(1.31, 1.37) | 23:21*<br>(23:08, 23:34) | 02:29*<br>(02:05, 02:53) | >3 hours | -1.7<br>(-2.1, -1.2) |
| *“drunk”* | 4.92<br>(4.65, 5.21) | 00:34*<br>(00:33, 00:36) | 01:25*<br>(01:23, 01:27) | <1 hour | 9.7<br>(9.3, 10) |
| *“dry eyes”* | 1.43<br>(1.39, 1.47) | 02:20*<br>(02:15, 02:24) | 03:50*<br>(03:42, 03:58) | 1-3 hours | 0.2<br>(0.11, 0.38) |
| *“glaucoma”* | 1.36<br>(1.33, 1.39) | 13:14*<br>(13:06, 13:22) | 01:05*<br>(23:39, 02:22) | >3 hours | -2.9<br>(-3, -2.7) |
| *“lasik”* | 1.18<br>(1.16, 1.21) | 11:13*<br>(10:55, 11:30) | 05:52*<br>(05:23, 06:24) | >3 hours | -1.5<br>(-1.7, -1.2) |
| *“pink eye”* | 1.77<br>(1.73, 1.80) | 04:37*<br>(04:35, 04:38) | 05:37*<br>(05:33, 05:40) | <1 hour | -0.32<br>(-0.67, 0.04) |
| *“visine”* | 1.35<br>(1.28, 1.42) | 00:58*<br>(00:43, 01:13) | 01:03*<br>(00:39, 01:26) | <1 hour | 0.43<br>(0.30, 0.55) |
| *“watery eyes”* | 1.41<br>(1.36, 1.46) | 01:54*<br>(01:47, 01:60) | 03:49*<br>(03:36, 04:02) | 1-3 hours | 0.26<br>(0.20, 0.31) |

## Discussion

Online search behavior patterns for terms related to common eye conditions and treatments exhibit diurnal variation that differed between terms, or for a single term by day of week. Additionally, although a limitation of our study is that we cannot control for non-clinical impacts on search behavior, several patterns reflected clinical understanding. For example, the observed increase in RSV from late night to early morning for “dry eyes” and “blurry eyes” is consistent with clinical reports of symptoms^4,7^. Stronger amplitude observed on weekends might represent elevated exposure to smoke or alcohol, which can increase symptoms^5,6^. The observed increase in RSV in from evenings to early mornings for “blurry eye”, “contact lenses”, “visine” and “watery eye” could reflect increased evening and nighttime symptoms of contact lens wearers^8^. Similarly, observed increases in RSV in mornings for “conjunctivitis” and “pink eye” may reflect clinical findings as well^1^. In comparison, diurnal search for “glaucoma”, “lasik” and “cataracts” occurred more during weekdays at daytimes (perhaps reflecting searches to schedule eye doctor appointments or obtain eye medications). This suggests information seeking behavior related to ocular procedures or chronic conditions not associated with acute symptoms may be more likely to occur during the regular work day.

## Conclusion

We have evidence from online hourly search patterns to suggest there are distinct times of day, night and week when online information seeking behavior for different eye conditions occurs. Precise temporal understanding of clinical eye disease presentation may be improved through future study using complementary data sources such as online search. This may also lead to improved diurnal eye disease monitoring or timing of resource allotment for telemedicine, healthcare messaging, and clinical interventions.

## Data Availability

All data is publicly available from Google Trends (https://trends.google.com/trends)

